# A Causal Inference of the Effect of Vaccination on COVID-19 Disease Severity and Need for Intensive Care Unit Admission Among Hospitalized Patients in an African Setting

**DOI:** 10.1101/2023.08.22.23294414

**Authors:** Eskedar Kebede Belayneh, Tigist Workneh Leulseged, Blen Solomon Teklu, Bersabel Hilawi Tewodros, Muluken Zeleke Megiso, Edengenet Solomon Weldesenbet, Mefthe Fikru Berhanu, Yohannes Shiferaw Shaweno, Kirubel Tesfaye Hailu

**Affiliations:** Department of Internal Medicine, MuluG Health Services, Addis Ababa, Ethiopia; Medical Research Lounge (Research Consultancy and Training Center), Addis Ababa, Ethiopia; Atlas College of Health Sciences, Addis Ababa, Ethiopia; University Medical Care, Silver Spring, Maryland, United States; Sheba medical center, Tel Aviv, Israel; Wachemo University, Hosanna, Ethiopia; Clinical Research Coordinator, Prolato Clinical Research Center, United States; Department of ENT, Adama General Hospital and Medical College, Adama, Ethiopia; Lecturer at Life Map higher learning Institute, Addis Ababa, Ethiopia

**Keywords:** COVID-19, Vaccination, Retrospective Cohort, Causal Inference, Ethiopia

## Abstract

**Background:** The COVID-19 pandemic is a respiratory illness that has spread to over 210 countries and killed over 6 million people. There is no specific treatment for COVID-19, but vaccines have been developed that can help prevent severe illness and death. A number of studies have investigated the effect of vaccination on disease severity and outcome, and the findings indicate that vaccination is linked to a significant reduction in the risk of hospitalization, intensive care unit admission, and death from the disease. However, there is a scarcity of evidence in Africa in general, and no similar study has been conducted in Ethiopia yet. Therefore, the study aimed to assess the effect of vaccination on COVID-19 disease severity and need for Intensive Care Unit (ICU) admission among hospitalized patients at a private specialty clinic in Ethiopia.

**Methods:** A retrospective cohort study was conducted among 126 patients with COVID-19, 41 vaccinated and 85 unvaccinated, who were hospitalized between September 2021 and May 2022. Data was summarized using frequency (percentage) and median (interquartile range). To compare the characteristics of the two groups, Chi-square/ Fisher’s Exact and Mann Whitney U tests at p-value of ≤ 0.05 were used. To identify the effect of vaccination on COVID-19 disease severity, Marginal Structural Model (MSM) with inverse probability weighting (IPW) approach using robust poisson regression model was fitted and adjusted relative risk (ARR) and 95% CI for ARR were used for interpreting the result.

**Results:** The cohort included groups that were fairly comparable in terms of their sociodemographic and clinical characteristics. More than half of the participants were older than 60 years (52.4%), were males (56.3%) and had one or more comorbid illness (52.4%). At admission 85 (67.5%) had severe disease and 11 (8.7%) progressed after hospitalization and required ICU admission, of which three unvaccinated cases died. From the final model, vaccination was found to be associated with a 62% decreased risk of developing severe COVID-19 disease if infected, compared to not getting vaccinated (ARR=0.38, 95% CI=0.23-0.65, p<0.0001).

**Conclusions:** The study’s findings support previous reports that vaccinated people are less likely to develop severe COVID-19 disease if infected with the virus, emphasizing the importance of continuing efforts to promote COVID-19 vaccination not only to safeguard individuals but also to confer community-level immunity.

## BACKGROUND

COVID-19 is a respiratory illness caused by a virus called severe acute respiratory syndrome coronavirus 2 (SARS-CoV-2). The virus was first identified in Wuhan, China, in December 2019. Since then, it has spread to over 210 countries and territories, infecting over 700 million people and killing over 6 million. And in Ethiopia, over 500 thousand confirmed cases and over 7 thousand deaths have been reported so far (1).

There is no specific treatment for COVID-19. Instead, supportive care is the mainstay of treatment, which includes measures such as rest, fluids, and medications to relieve symptoms. In severe cases, patients may need oxygen therapy or mechanical ventilation (2-4). As a more effective strategy, a vaccine to prevent COVID-19 was developed. The first COVID-19 vaccine was approved by the U.S. Food and Drug Administration on August 23, 2021 (5). Since then, millions of people around the world have been vaccinated. As of August 9, 2023, a total of 13,492,264,486 vaccine doses have been administered globally (1). The Ethiopian Federal Ministry of Health launched COVID-19 immunization on March 13, 2021, and as of May 27, 2023, a total of 68,856,793 vaccine doses have been provided (1,6).

While the safety and efficacy of the vaccines have been proven in controlled studies and are expected to reduce morbidity and mortality, there is an irrefutable need for further evidence of the effectiveness of the vaccines in preventing severe illness and death from COVID-19 in the real world (7-21). With this in mind, a number of studies have investigated the effect of COVID-19 vaccination on disease severity and outcome. According to these studies, Vaccination has been linked to a significant reduction in the risk of hospitalization, intensive care unit admission, and death from COVID-19 (22-30). However, there is a scarcity of evidence throughout Africa in general, and in Ethiopia in particular (31-32). Moreover, disease epidemiology in Africa appears to differ from that of the rest of the world thus far (33-40). Given these disparities in disease epidemiology and the lack of adequate evidence, it is especially important to understand the potential changes in disease patterns in the post-vaccination era in a local setting. Findings from such research can be used to make informed decisions at both the public health policy design and individual levels. Therefore, the aim of this study was to assess the effect of vaccination on COVID-19 disease severity and need for ICU admission among hospitalized patients at a private specialty clinic in Ethiopia from September 2021 to May 2022.

## METHODS

### Study Setting and Design

An institution-based retrospective cohort study, following the STROBE checklist for cohort studies, was conducted from June to August, 2023 among COVID-19 patients who were admitted at a private clinic, MuluG health Services, between September 2021 and May 2022. The hospital is a primary Internal Medicine, Gynecology and pediatric specialty center, which had a dedicated ward for COVID-19 patients management during the pandemic. The cohort was classified into two groups based on their vaccination status for COVID-19 as fully/partially vaccinated (exposed group) and unvaccinated (non-exposed group).

### Population and Sample size

The study included all laboratory-confirmed COVID-19 patients who were admitted and completed their follow-up at the hospital between September 2021 and May 2022 and had complete data on basic clinical and laboratory parameters, and COVID-19 vaccination status. Accordingly, from the 184 admissions, 126 eligible participants (41 vaccinated and 85 unvaccinated) were included in the study.

A post-hoc power analysis was calculated using G*Power 3.19.4 to check the power of the study using a two-tailed z-test for difference between two independent proportions with the following statistical parameters; 5% level of significance, proportion of severe COVID-19 and sample size in the vaccinated group of 36.6% and 41, respectively; and proportion of severe COVID-19 and sample size in the non-vaccinated group of 82.4% and 85, respectively. Finally, the power of the study was found to be 99.9%.

### Operational Definitions

- **Moderate COVID-19 Disease:** Individuals who show evidence of lower respiratory disease during clinical assessment or imaging and who can maintain oxygenation in room air (2, 4).
- **Severe COVID-19 Disease:** Includes patients who have SPO2 ≤ 93% on atmospheric air or a ratio of arterial partial pressure of oxygen to fraction of inspired oxygen (PaO2/FiO2) (SF ratio < 315), in respiratory distress or respiratory rate >30 breaths/minutes, or lung infiltrates involving >50% of the chest (2, 4).

### Data Collection Procedures and Quality Assurance

Data on socio-demographic, clinical, laboratory and vaccination status of the patients was extracted from the electronic data registry system that was designed using the WHO CRF patient management and follow-up form. Data quality was assured through proper training of two data collectors (General Practitioners) on the tool, double data entry, and data cleaning through checking for inconsistencies, numerical errors and missing parameters. All data management and analysis were performed using STATA software version 17.0 (College Station, TX).

### Statistical Analysis

The study population characteristics was summarized and presented using frequency with percentage for categorical variables. Symptom duration, measured in days, was summarized using median with interquartile range (IQR) due to the skewed distribution of the data (Kolmogorov-Smirnov and Shapiro-Wilk tests p-value <0.0001). To compare the socio-demographic and clinical characteristics within the cohort (Vaccinated Vs Unvaccinated), a Chisquare test was used. Whenever the assumption of a chi-square test where no cell should have expected count of <5 failed, Fisher’s exact test was used instead. To compare the median symptom duration between the groups, a Mann Whitney U test was used. For all tests, a statistically significant difference was detected for variables with a p-value of ≤ 0.05.

To identify the effect of vaccination on COVID-19 disease severity (moderate vs severe), Marginal Structural Model (MSM) with inverse probability weighting (IPW) approach was used in two steps.

First, the treatment model using a binary logistic regression model was run to estimate the probability of vaccination status given the covariates (propensity score). The estimated probability of vaccination status was then used to compute the inverse probability weights for each individual. The inverse of the probability of vaccination was then used to weight each individual in the estimation of the marginal odds ratio. Univariate analysis at 25% level of significance and clinical judgment was used to select variables to be included in the treatment model.

Subsequently, the final outcome model, using the vaccination status variable adjusted for inverse probability weights, was fitted to predict COVID-19 disease severity. To identify the effect of vaccination on disease severity, a Robust Poisson regression model was selected. Fitness of the model was assessed using Pearson chi-square and Deviance tests and the model fit the data well. Accordingly, on the final model, a p-value of ≤ 0.05 indicates that vaccination is a significant predictor of COVID-19 disease severity. In case of significant relationship, adjusted relative risk (ARR) and 95% CI for ARR were used for interpreting the result. Effect of vaccination on need for ICU admission was not assessed due to the small frequency of the outcome.

## RESULTS

### Socio-demographic and Clinical Characteristics

More than half of the participants were older than 60 years (52.4%) and were males (56.3%). The frequent presenting symptom was cough in 119 (94.4%) followed by anorexia in 96 (76.2%), and constitutional symptoms of myalgia in 63 (50.0%), arthralgia in 62 (49.2%), and headache in 51 (40.5%). Anosmia and/or ageusia were reported by 58 (46.0%). The median duration of symptoms was 7 days (IQR, 4-10 days), with a minimum of 1 day and a maximum of 20 days. Sixty-six patients (52.4%) presented with one or more comorbid conditions, with hypertension and diabetes being the most prevalent, affecting 38 patients (30.2%) each. .

According to the Chi-square/Fisher’s exact and Mann Whitney U tests, there was no statistically significant difference in age, sex, presenting symptom, symptom duration or comorbid illness history between vaccinated and unvaccinated patients. (**Table 1**)

**Table 1:** Socio–demographic and clinical characteristics of COVID-19 patients in Ethiopia (n=126)

| Variable |  | Total (%) | Took vaccine |  | p-value |
| --- | --- | --- | --- | --- | --- |
|  |  |  | No (%) | Yes (%) |  |
| Age (in years) | ≤ 40 | 14 (11.1) | 11 (12.9) | 3 (7.3) | 0.238 |
|  | 40-60 | 46 (36.5) | 34 (40.0) | 12 (29.3) |  |
|  | ≥ 60 | 66 (52.4) | 40 (47.1) | 26 (63.4) |  |
| Sex | Female | 55 (43.7) | 33 (38.8) | 22 (53.7) | 0.116 |
|  | Male | 71 (56.3) | 52 (61.2) | 19 (46.3) |  |
| Cough | No | 7 (5.6) | 4 (4.7) | 3 (7.3) | 0.681 |
|  | Yes | 119 (94.4) | 81 (95.3) | 38 (92.7) |  |
| Shortness of breath | No | 84 (66.7) | 60 (70.6) | 24 (58.5) | 0.179 |
|  | Yes | 42 (33.3) | 25 (29.4) | 17 (41.5) |  |
| Chest pain | No | 108 (85.7) | 75 (88.2) | 33 (80.5) | 0.244 |
|  | Yes | 18 (14.3) | 10 (11.8) | 8 (19.5) |  |
| Fever | No | 82 (65.1) | 54 (63.5) | 28 (68.3) | 0.599 |
|  | Yes | 44 (34.9) | 31 (36.5) | 13 (31.7) |  |
| Headache | No | 75 (59.5) | 52 (61.2) | 23 (56.1) | 0.586 |
|  | Yes | 51 (40.5) | 33 (38.8) | 18 (43.9) |  |
| Arthralgia | No | 64 (50.8) | 47 (55.3) | 17 (41.5) | 0.146 |
|  | Yes | 62 (49.2) | 38 (44.7) | 24 (58.5) |  |
| Myalgia | No | 63 (50.0) | 45 (52.9) | 18 (43.9) | 0.342 |
|  | Yes | 63 (50.0) | 40 (47.1) | 23 (56.1) |  |
| Anorexia | No | 30 (23.8) | 21 (24.7) | 9 (22.0) | 0.734 |
|  | Yes | 96 (76.2) | 64 (75.3) | 32 (78.0) |  |
| Anosmia/ageusia | No | 68 (54.0) | 49 (57.6) | 19 (46.3) | 0.233 |
|  | Yes | 58 (46.0) | 36 (42.4) | 22 (53.7) |  |
| Diarrhea | No | 115 (91.3) | 76 (89.4) | 39 (95.1) | 0.501 |
|  | Yes | 11 (8.7) | 9 (10.6) | 2 (4.9) |  |
| vomiting | No | 116 (92.1) | 79 (92.9) | 37 (90.2) | 0.727 |
|  | Yes | 10 (7.9) | 6 (7.1) | 4 (9.8) |  |
| Nausea | No | 113 (89.7) | 79 (92.9) | 34 (82.9) | 0.117 |
|  | Yes | 13 (10.3) | 6 (7.1) | 7 (17.1) |  |
| Symptom duration (Median, IQR) |  | 7.0 (4.0-10.0) | 7.0 (4.5-10.0) | 7.0 (4.0-9.0) | 0.619 |
| Diabetes | No | 88 (69.8) | 57 (67.1) | 31 (75.6) | 0.327 |
|  | Yes | 38 (30.2) | 28 (32.9) | 10 (24.4) |  |
| Hypertension | No | 88 (69.8) | 60 (70.6) | 28 (68.3) | 0.793 |
|  | Yes | 38 (30.2) | 25 (29.4) | 13 (31.7) |  |
| HIV | No | 120 (95.2) | 82 (96.5) | 38 (92.7) | 0.390 |
|  | Yes | 6 (4.8) | 3 (3.5) | 3 (7.3) |  |
| Asthma | No | 123 (97.6) | 82 (96.5) | 41 (100.0) | 0.550 |
|  | Yes | 3 (2.4) | 3 (3.5) | 0 |  |
| Cardiac disease | No | 120 (95.2) | 82 (96.5) | 38 (92.7) | 0.390 |
|  | Yes | 6 (4.8) | 3 (3.5) | 3 (7.3) |  |

### Vital sign and Laboratory Parameters

Upon baseline assessment, only a smaller proportion of patients presented with abnormal vital signs and laboratory parameters. Of these, 21.4% had elevated systolic blood pressure (SBP), 14.3% had elevated diastolic blood pressure (DBP), 32.5% had an elevated pulse rate (PR), and 7.1% had hyperthermia. Furthermore, 27.8% had an elevated neutrophil to lymphocyte ratio (NLR), 57.9% had an elevated hematocrit, 27.0% had a platelet count of less than 150 x 103/ul or greater than 450 x 103/ul, 23.0% had an elevated blood urea nitrogen (BUN) level, and 31.7% had an elevated creatinine level. On the other hand, the majority (71.4%) presented with hypothermia.

Additionally, the result showed that unvaccinated patients were more likely to have a raised pulse rate (38.8% Vs 19.5%, p-value=0.030), and BUN (28.2% Vs 12.2%, p-value=0.045) than vaccinated patients. Otherwise, there were no significant differences between the two groups in terms of their blood pressure, oxygen saturation, temperature, NLR, hematocrit level, platelet count, or creatinine level. (**Table 2**)

**Table 2:** Vital sign and laboratory parameters of COVID-19 patients in Ethiopia (n=126)

| Variable |  | Total (%) | Took COVID vaccine |  | p-value |
| --- | --- | --- | --- | --- | --- |
|  |  |  | No (%) | Yes (%) |  |
| SBP (mmHg) | < 140 | 99 (78.6) | 70 (82.4) | 29 (70.7) | 0.136 |
|  | ≥ 140 | 27 (21.4) | 15 (17.6) | 12 (29.3) |  |
| DBP (mmHg) | < 90 | 108 (85.7) | 74 (87.1) | 34 (82.9) | 0.535 |
|  | ≥ 90 | 18 (14.3) | 11 (12.9) | 7 (17.1) |  |
| PR (per minute) | <100 | 85 (67.5) | 52 (61.2) | 33 (80.5) | **0.030\*** |
|  | ≥ 100 | 41 (32.5) | 33 (38.8) | 8 (19.5) |  |
| Temperature (°C) | < 36.5 | 90 (71.4) | 62 (72.9) | 28 (68.3) | 0.653 |
|  | 36.5-37.5 | 27 (21.4) | 18 (21.2) | 9 (22.0) |  |
|  | > 37.5 | 9 (7.1) | 5 (5.9) | 4 (9.8) |  |
| NLR | ≤ 3 | 37 (29.4) | 26 (30.6) | 11 (26.8) | 0.657 |
|  | 3-6 | 35 (27.8) | 22 (25.9) | 13 (31.7) |  |
|  | 6-9 | 26 (20.6) | 16 (18.8) | 10 (24.4) |  |
|  | >9 | 28 (22.2) | 21 (24.7) | 7 (17.1) |  |
| Hematocrit (%) | ≤ 45 | 53 (42.1) | 35 (41.2) | 18 (43.9) | 0.772 |
|  | >45 | 73 (57.9) | 50 (58.8) | 23 (56.1) |  |
| Platelet (x 10<sup>3</sup>/ul) | 150-450 | 92 (73.0) | 60 (70.6) | 32 (78.0) | 0.377 |
|  | <150/>450 | 34 (27.0) | 25 (29.4) | 9 (22.0) |  |
| BUN (mg/dl) | < 20 | 97 (77.0) | 61 (71.8) | 36 (87.8) | **0.045\*** |
|  | ≥ 20 | 29 (23.0) | 24 (28.2) | 5 (12.2) |  |
| Creatinine (mg/dl) | <1.1 | 86 (68.3) | 59 (69.4) | 27 (65.9) | 0.688 |
| | $\geq 1.1$ | 40 (31.7) | 26 (30.6) | 14 (34.1) | |

**Note:** \*statistically significant

### COVID-19 Disease Severity and Need for ICU Admission

Of the 126 hospitalized COVID-19 patients, 85 (67.5%, 95% CI=59.7-76.8%) had severe disease, while 41 (32.5%, 95% CI= 23.2-40.3%) had moderate disease upon admission.

Disease progression after hospitalization that necessitated ICU care occurred in 11 (8.7%, 95%=4.9-15.2%). One of the 11 cases was vaccinated and was later discharged improved. The remaining ten were unvaccinated, of which seven were discharged improved and three died.

### Treatment model: Logistic regression of factors associated with COVID-19 vaccination

The treatment model using a binary logistic regression model was run by including the following categories: age category, sex, fever, headache, arthralgia, anorexia, anosmia/ageusia, diarrhea, vomiting, nausea, diabetes, hypertension, SBP, DBP, PR, temperature, NLR, hematocrit, platelet, Bun, and creatinine. By fitting the final treatment model, propensity score was estimated and it was used to compute the inverse probability weights for each individual. The inverse of the probability of vaccination status was then used to weight each individual in the estimation of the marginal odds ratio. (**Table 3**)

**Table 3:** Binary Logistic Regression model of factors associated with vaccination status among COVID-19 patients in Ethiopia (n=126)

| Variable | AOR | 95% CI AOR | p-value |
| --- | --- | --- | --- |
| **Age in years (R: $\leq 40$)** |  |  |  |
| 40-60 | 1.45 | 0.20, 10.51 | 0.715 |
| $\geq 60$ | 3.16 | 0.40, 24.85 | 0.275 |
| **Sex (Male)** | 0.58 | 0.20, 1.69 | 0.316 |
| **Fever (Yes)** | 0.94 | 0.26, 3.36 | 0.924 |
| **Headache (Yes)** | 1.68 | 0.56, 5.06 | 0.356 |
| **Arthralgia (Yes)** | 2.57 | 0.64, 10.32 | 0.182 |
| **Myalgia (Yes)** | 0.65 | 0.16, 2.66 | 0.552 |
| **Anorexia (Yes)** | 0.29 | 0.07, 1.15 | 0.078 |
| **Anosmia/ageusia (Yes)** | 1.73 | 0.52, 5.74 | 0.367 |
| **Diarrhea (Yes)** | 0.12 | 0.01, 1.28 | 0.080 |
| **Vomiting (Yes)** | 1.00 | 0.11, 9.27 | 0.998 |
| **Nausea (Yes)** | 5.49 | 0.88, 34.14 | 0.068 |
| **Diabetes (Yes)** | 0.76 | 0.25, 2.28 | 0.624 |
| **Hypertension (Yes)** | 0.93 | 0.28, 3.11 | 0.902 |
| **SBP ($\geq 140$ mmHg)** | 3.55 | 0.89, 14.15 | 0.072 |
| **DBP ($\geq 90$ mmHg)** | 0.89 | 0.17, 4.69 | 0.888 |
| **PR** ($\geq 100$ /minute) | 0.24 | 0.07, 0.77 | 0.017 |
| **Temperature** (R: $<36.5$ oC) | | | |
| 36.5-37.5 oC | 2.61 | 0.73, 9.31 | 0.140 |
| $>37.5$ oC | 2.84 | 0.31, 25.94 | 0.355 |
| **NLR** (R: $\leq 3.00$) | | | |
| 3.01-5.99 | 1.22 | 0.32, 4.69 | 0.772 |
| 6.00-9.00 | 1.23 | 0.29, 5.19 | 0.774 |
| $\geq 9.01$ | 0.58 | 0.11, 3.18 | 0.532 |
| **Hematocrit** ($> 45\%$) | 1.40 | 0.47, 4.18 | 0.543 |
| **Platelet** ($<150/>450 \times 10^3/\text{ul}$) | 0.64 | 0.21, 1.97 | 0.438 |
| **BUN** ($\geq 20$ mg/dl) | 0.20 | 0.04, 0.90 | 0.036 |
| **Creatinine** ($\geq 1.1$ mg/dl) | 2.22 | 0.63, 7.84 | 0.215 |

**Note:** AOR, Adjusted Odds ratio; CI, Confidence interval; \*statistically significant

### Outcome Model: Effect of vaccination on COVID-19 disease severity

To identify the effect of vaccination on COVID-19 disease severity, a Robust Poisson regression model was run by fitting the vaccination status variable adjusted for inverse probability weights, that was obtained from the treatment model above.

The result shows that vaccination is associated with a 62% decreased risk of developing severe COVID-19 disease if infected, compared to not getting vaccinated **(ARR=0.38, 95% CI=0.23-0.65, p<0.0001)**. (**Table 4**)

**Table 4:** Robust poisson regression of effect of vaccination on disease severity among COVID-19 patients in Ethiopia (n=126)

|  | COVID-19 Severity |  | ARR | 95% CI for ARR | P-value |
| --- | --- | --- | --- | --- | --- |
|  | Moderate | Severe |  |  |  |
| **Vaccination** |  |  |  |  |  |
| No | 15 | 70 | 1 | 1 |  |
| Yes | 26 | 15 | **0.38** | **0.23, 0.65** | **$<0.0001^*$** |

**Note:** ARR, Adjusted Relative Risk; CI, Confidence interval; \*statistically significant

## DISCUSSION

This retrospective cohort study investigated the effect of vaccination on COVID-19 disease severity among hospitalized patients who were on follow-up at a private specialty clinic in Ethiopia between September 2021 and May 2022. Although multiple studies are done internationally, this is the first of its kind to answer this research question at an Ethiopian institution. The study included 126 hospitalized COVID-19 patients, 41 vaccinated and 85 unvaccinated. The participants were fairly comparable in terms of their socio-demographic and clinical characteristics. More than half of the participants were older than 60 years (52.4%), were males (56.3%) and had one or more comorbid illness (52.4%). At admission 85 (67.5%) had severe disease and 11 (8.7%) progressed after hospitalization and required ICU admission, of which three unvaccinated cases died.

According to the Robust Poisson regression analysis using the MSM model with the IPW approach, vaccination is causally associated with disease severity and progression in a protective way, with vaccinated people 62% less likely to develop severe COVID-19 disease if infected with the virus. The findings indicate that immunization plays a significant role in preventing the development of severe disease. This is a noteworthy finding, as it suggests that vaccination can help to protect people from the most serious consequences of COVID-19; hospitalization, longterm complications, and even death. Furthermore, it also contributes to the greater goal of reducing overall healthcare costs and preventing the spread of illness. The study’s findings are consistent with the results of other studies that have shown that vaccination is effective in preventing severe COVID-19 in both Africa (31-32) and outside Africa (22-30). Although the study findings note the importance of vaccination in reducing morbidity and mortality, a significant portion of the population in Ethiopia is unvaccinated (1,6). Hence, the importance of vaccination in reducing morbidity and mortality in combination with other preventative measures is a crucial instrument in the fight against COVID-19 and should not be overlooked by the general public.

When interpreting the study findings, bear the following strengths and limitations in mind. The study’s strengths include the fact that it is the first in Ethiopia and one of only a handful in Africa, it was conducted in a hospital setting where a more complete picture of the influence of vaccine on those who are already infected and required admission can be studied, and the causal effect of vaccination was estimated using a causal inference model which can provide strong evidence of causality from observational data. Its limitations are that further investigation of the vaccine effect in terms of type, dose, and period between vaccination and initial symptom could not be made since they were not documented, and the generalizability is limited as it is a single center study.

## CONCLUSION

The study shows that vaccinated people are less likely to develop severe COVID-19 disease if infected with the virus. Its findings are consistent with studies done to answer similar questions during the clinical trials for COVID-19 vaccine development and post-market surveillance, emphasizing the importance of continuing efforts to promote COVID-19 vaccination not only to safeguard individuals but also to confer community-level immunity. We recommend large prospective community and multicenter studies to gain a better understanding of the effect of vaccination on short- and long-term disease outcomes, taking into account the dose and type of vaccine.

## Data Availability

All relevant data are available upon reasonable request from Tigist W. Leulseged.

## Declaration

### Ethics approval and consent to participate

The study was conducted after obtaining ethical clearance from Addis Ababa Regional Health Bureau Ethics Review Committee (AARHB-ERC). The AARHB-ERC also waived the need for informed consent as the study used secondary data. Anonymity of the participants was maintained by use of medical record number in the research report. No other personal identifiers of the patients were used in the research report. Access to the collected information was limited to the investigators and confidentiality was maintained throughout the project.

### Consent for publication

Not applicable

### Availability of data and materials

All relevant data are available upon reasonable request from Tigist W. Leulseged.

### Authors’ Contribution

EKB and TWL conceived and designed the study. BST, BHT, MZM, ESW, MFB, YSS, and KTH contributed to the conception and design of the study. TWL performed statistical analysis, and drafted the initial manuscript. BST, BHT, MZM, and KTH contributed to the statistical analysis and interpretation of the findings. EKB, ESW, MFB, and YSS revised the manuscript. All authors approved the final version of the manuscript.

## Acknowledgment

The authors would like to thank all individuals involved in the facilitation of extraction of the electronic data.

